# Household transmission of SARS-CoV-2: a systematic review and meta-analysis of secondary attack rate

**DOI:** 10.1101/2020.07.29.20164590

**Authors:** Zachary J. Madewell, Yang Yang, Ira M. Longini, M. Elizabeth Halloran, Natalie E. Dean

**Author notes:** Correspondence to: Zachary J. Madewell, Department of Biostatistics, University of Florida, PO Box 117450, Gainesville, FL 32611.

## Abstract

**Background:** Severe acute respiratory syndrome coronavirus 2 (SARS-CoV-2) is spread by direct, indirect, or close contact with infected people via infected respiratory droplets or saliva. Crowded indoor environments with sustained close contact and conversations are a particularly high-risk setting.

**Methods:** We performed a meta-analysis through July 29, 2020 of SARS-CoV-2 household secondary attack rate (SAR), disaggregating by several covariates (contact type, symptom status, adult/child contacts, contact sex, relationship to index case, index case sex, number of contacts in household, coronavirus).

**Findings:** We identified 40 relevant published studies that report household secondary transmission. The estimated overall household SAR was 18·8% (95% confidence interval [CI]: 15·4%–22·2%), which is higher than previously observed SARs for SARS-CoV and MERS-CoV. We observed that household SARs were significantly higher from symptomatic index cases than asymptomatic index cases, to adult contacts than children contacts, to spouses than other family contacts, and in households with one contact than households with three or more contacts.

**Interpretation:** To prevent the spread of SARS-CoV-2, people are being asked to stay at home worldwide. With suspected or confirmed infections referred to isolate at home, household transmission will continue to be a significant source of transmission.

## Introduction

The coronavirus disease 2019 (COVID-19) pandemic is caused by severe acute respiratory syndrome coronavirus 2 (SARS-CoV-2). First identified in Wuhan, China, in January 2020, SARS-CoV-2 has now been reported worldwide in 214 countries and territories.^1^ Although COVID-19 often presents clinically as a mild disease, it may cause severe illness or even death, particularly among older individuals and those with concurrent chronic diseases.^2,3^ SARS-CoV-2 is spread via direct, indirect, or close contact with infected people via infected respiratory droplets or saliva.^4^ Airborne and fomite transmission are other potential routes.^5^ Crowded indoor environments with sustained close contact and conversations are a particularly high-risk setting.^6^

Stay-at-home orders implemented in response to the pandemic reduced human mobility by 35– 63% in the USA,^7^ 63% in the UK,^8^ and 54% in Wuhan,^9^ relative to normal conditions. This concomitantly increased time spent at home and likely increased household transmission of SARS-CoV-2. For example, following campaigns promoting social distancing and bans on social gatherings, Iceland observed a shift in exposure from international travel and social exposure to exposure in the household environment.^10^ The WHO-China Joint Mission reported that most locally generated cases were clustered in households.^11^ While current CDC recommendations are to maintain six feet distance from when a household member is sick, this may be difficult to achieve in practice nor be fully effective.^12^

Studies of household contacts are uniquely advantageous for understanding transmission dynamics. Besides characterizing transmissibility in a household setting, household secondary attack rate (SAR) provides a useful estimate of both the susceptibility of contacts and infectiousness of index cases. Studies can collect detailed data on the type, timing and duration of contacts. Researchers can examine features of the household, such as density or sanitization methods.^13^ This information may be used to inform control measures.

We conducted a review of the rapidly growing body of literature describing household transmission of SARS-CoV-2. We describe the types of study designs available and then present a meta-analytic summary of transmission within households for SARS-CoV-2, disaggregated by several exposures. We discuss available evidence for asymptomatic and presymptomatic exposure, as well as correlates of susceptibility of household contacts and infectivity of index cases. We also explore how many households with index cases had any secondary transmission, and compare household transmission with other coronaviruses.

### Review of transmissibility of SARS-CoV-2 in households and families

We estimated transmissibility of SARS-CoV-2 within the household or family by the crude secondary attack rate (SAR), or the probability that an exposed susceptible person develops disease over the duration of infectiousness in a case patient. The denominator of the SAR is the number of exposed contacts, and the numerator is the number who become infected with SARS-CoV-2 or develop COVID-19.

To estimate household SAR, we searched PubMed using the terms “SARS-CoV-2” or “COVID-19” plus: “secondary attack rate,” “household,” “close contacts,” “contact transmission,” “contact attack rate,” “family transmission,” or “family attack rate.” We extracted all articles with original data for estimating household SAR of SARS-CoV-2. The publication must report a numerator and denominator among household contacts, or at least two of numerator, denominator, and SAR. Where numerators (numbers of infected contacts) or denominators (numbers of contacts) were not reported but the number of index cases and SAR were available, the denominator was calculated acknowledging limits of significant digits. The last search was conducted on July 29, 2020.

We identified 40 relevant published studies that report household secondary transmission (Figure 1; S1 Table). Most studies were in China,^14-32^ but others were in South Korea,^33-37^ USA,^38-41^ Spain,^42,43^ Australia,^44^ Brazil,^45^ Brunei,^46^ Germany,^47^ India,^48^ Israel,^49^ Italy,^50^ Singapore,^51^ Taiwan,^52^ and UK.^53^

**Figure 1.**
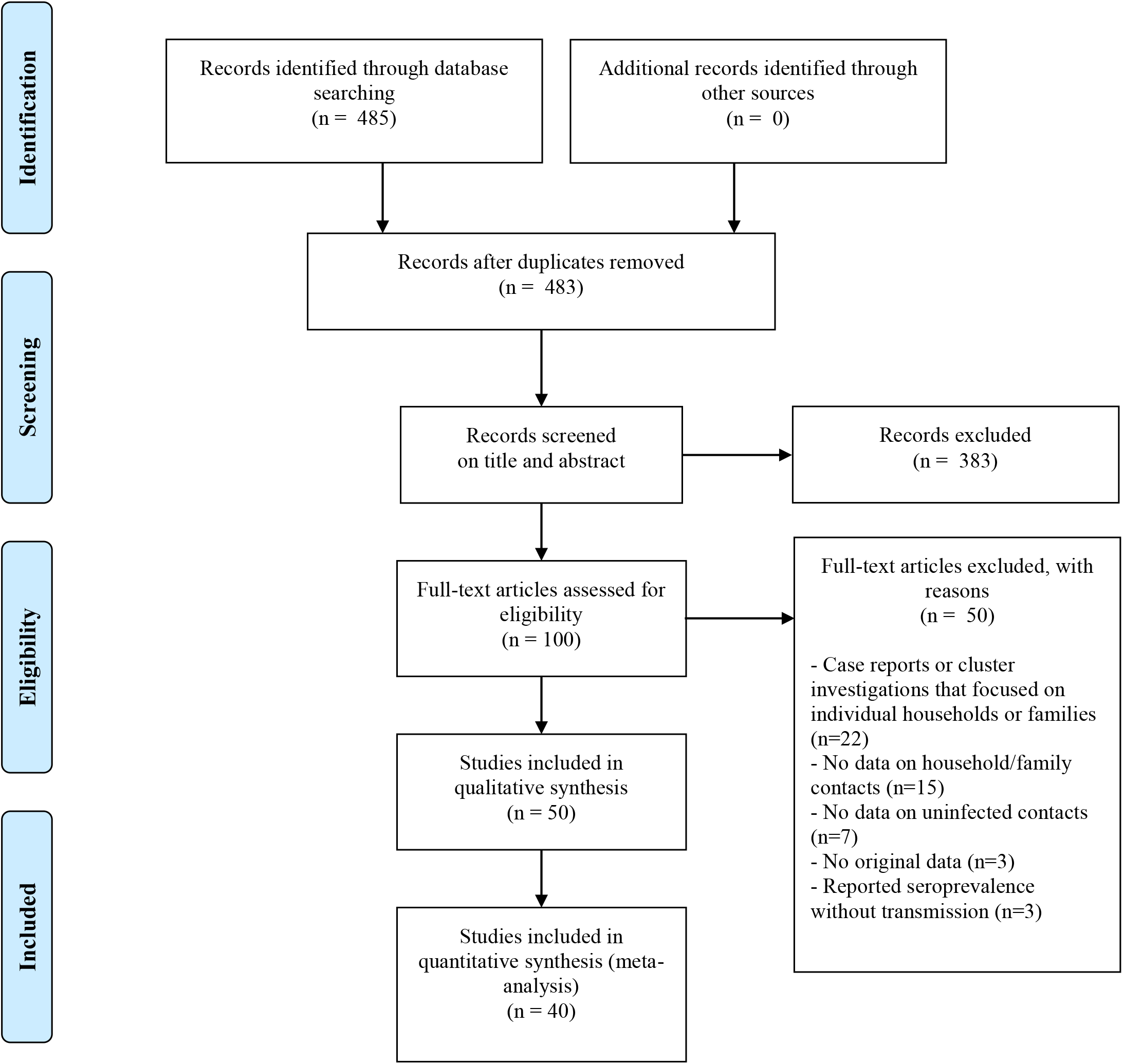
Flowchart for review of household secondary attack rate

Much of our understanding of household transmission is derived from contact tracing investigations. After a household member is confirmed to be infected with SARS-CoV-2, household or family contacts are identified, followed for a short duration of time (e.g., two weeks), and tested if eligible for testing according to national testing guidelines to ascertain secondary infections. Index cases were identified by passive surveillance,^21,28,29,31,38-40,43,44,49-51^ active surveillance of key populations (e.g., travelers from areas with active SARS-CoV-2 transmission, and individuals detected by neighborhood fever screenings),^25,27,34^ both active and passive surveillance,^14,15,17-20,22,24,26,30,32,33,35,36,41,46,48,52,53^ or regional seroprevalence surveys.^42,45^ We did not include case reports or cluster investigations that focused on individual households or families in an effort to reduce publication bias.

Some studies included index cases with SARS-CoV-2 infections (both symptomatic and asymptomatic),^14,15,18-22,24-27,32-34,36,37,41,42,47,49,52^ whereas others included symptomatic COVID-19 index cases only.^16,28-31,38-40,43,44,50,51,53^ Another targeted asymptomatic SARS-CoV-2 infected index cases,^35^ some of whom developed symptoms during a follow-up period. Most studies did not describe how co-primary index cases were handled or whether secondary infections could have been acquired from outside the household, both of which can inflate the crude SAR. Several stated they assumed all secondary cases were infected by the index case to whom they were traced,^26,33,48^ another excluded secondary cases if they developed symptoms before exposure to the primary case,^52^ and another randomly selected one index case as the infector.^18^ Ignoring tertiary transmission also inflates the SAR.

Many studies only included household contacts,^14,26,28,29,31,35,37,38,40,41,43,49,50,53^ but others included family members,^15,20,21,23,27,30^ or other close contacts,^16-19,22,24,25,32-34,36,39,42,44,46-48,52^ including individuals outside the household. Of the latter studies of close contacts, many reported SARs among household contacts,^17,18,22,24,25,32-34,36,39,42,44,46-48,52^ or all family members.^16,19-21,23,27,30^ We did not include studies of close contacts that did not report SARs for household or family members. We assumed that studies of household contacts included anyone living in the same household as the index case unless stated otherwise. For example, several studies reported household contacts as family members in households.^25,28,32,45^ Several studies further restricted household contacts to those who spent at least one night or 24 hours in the house after symptom onset of the index case.^26,28,31^

Most studies involved tracing contacts and monitoring them for 14,^14,16-18,22,24,28-34,37-39,44,47,48,50,52^ or 21 days.^19,26^ Monitoring methods included phone calls,^21,34,39,44^ text messages,^39,44^ or direct observation by healthcare workers.^17,33^ Some studies tested all contacts immediately after the index case was diagnosed at the onset of the observation period and monitored them for symptoms.^18,35,40,46,51^ Several studies that reported testing all contacts irrespective of symptoms also reported extra testing of individuals who developed symptoms during quarantine.^25,31,38,40,46^ Others tested all contacts during or at the end of the observation period regardless of symptoms,^24,28,37,48^ whereas others only tested symptomatic contacts.^29,39,44,50,52^ Several studies tested contacts multiple times throughout the observation period irrespective of symptoms.^14,17,19,22,25,26,30,31,33,38,47^ Other studies tested all contacts,^15,23,40-42,45^ or interviewed index cases about symptoms of household members,^43,53^ immediately without additional monitoring. Many studies, particularly those in China, reported in-home quarantine of contacts during the observation period after index cases were confirmed.^14,17-19,22,24,29-34,36,37,44,46,47,50-52^

Case ascertainment was primarily done via RT-PCR on nasopharyngeal or oropharyngeal samples.^14,17,18,21-23,26-33,35-37,40,44,46-53^ However, several studies also reported whole-genome sequencing,^27,28,32,47^ nucleic acid tests,^19,25,27^ and antibody tests.^42,45^ Two studies identified index cases via RT-PCR, but only collected symptom information about household contacts from telephone interviews with index cases.^43,53^

### Secondary attack rate

Meta-analyses were done using a restricted maximum-likelihood estimator model to yield a point estimate and 95% confidence interval (CI) for SAR by exposure type. Each study was treated as a random effect and exposure (contact type, symptom status, adult/child contacts, contact sex, relationship to index case, index case sex, number of contacts in household, coronavirus) as a fixed effect. The Cochran Q-test is reported as a measure of heterogeneity. All tests of significance were at the α = 0·05 level. All analyses were done in R 4·0·2 using the package metafor.^54,55^

Figure 2 summarizes overall SARs for the 40 studies included. If a study reported SARs for both household and family contacts, we only included the household contact SAR to avoid counting the same individuals twice. The estimated mean SAR for household contacts is 19·0% (95% CI: 14·9%–23·1%) and family contacts is 18·1% (95% CI: 12·9%–34·8%), both with significant heterogeneity (*P* < 0·001). SARs were not significantly different between household and family contacts, but they were nearly five times higher than SARs reported for close contacts (4·3%; 95% CI: 2·9%–5·6%) (*P* < 0·001) (S1 Figure). Among studies that included other forms of contact outside the household, household contact was reported as a significant risk factor for infection.^17,18,24,25,32,33,46,48^ The household SAR for studies in China was not significantly different than SARs reported in studies from other countries (S2 Figure). There was no significant difference in SARs between studies that tested symptomatic contacts (17·8%; 95% CI: 2·1%–33·6%) versus all contacts (19·7%; 95% CI: 15·4%–24·0%), both with significant heterogeneity (*P* < 0·001) (S3 Figure).

**Figure 2.**
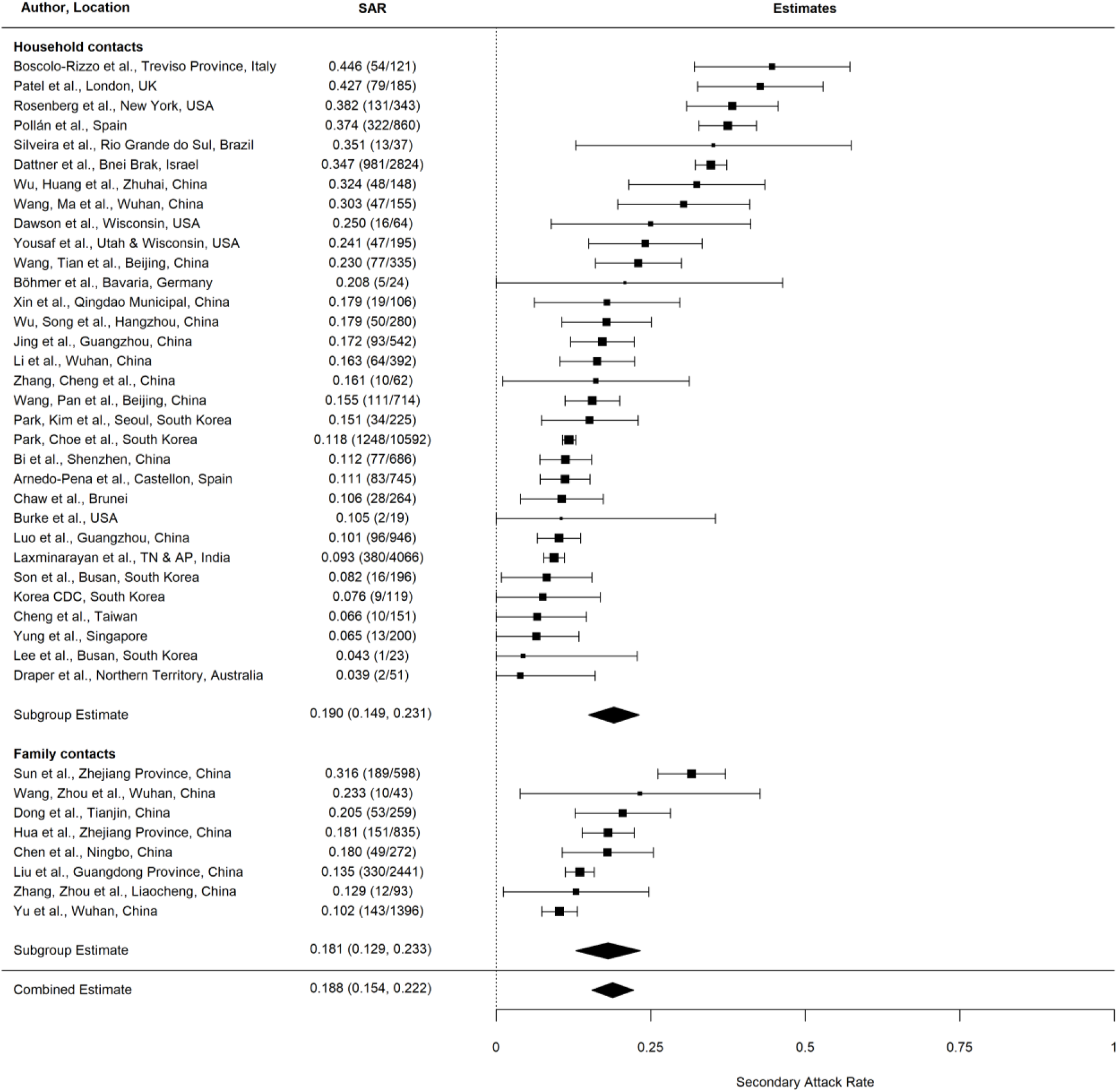
Secondary attack rates (SAR) for household contacts and family contacts (including individuals outside the index case household).

### Presymptomatic, asymptomatic, and symptomatic transmission

To study the transmissibility of asymptomatic SARS-CoV-2 index cases, Figure 3 summarizes 17 studies reporting household SARs from symptomatic index cases and four from asymptomatic index cases. The estimated mean household SAR from symptomatic index cases (19·9%; 95% CI: 14·0%– 25·7%) was significantly higher than from asymptomatic index cases (0·7%; 95% CI: 0%–3·8%) (*P* < 0·001), though there were only a few studies of asymptomatic index cases. These findings are consistent with other household studies that reported asymptomatic index cases as having limited or no role in household transmission,^17,25,52^ although many studies acknowledged that asymptomatic index cases are likely under-represented.^18,30,33,43,46,53^

**Figure 3.**
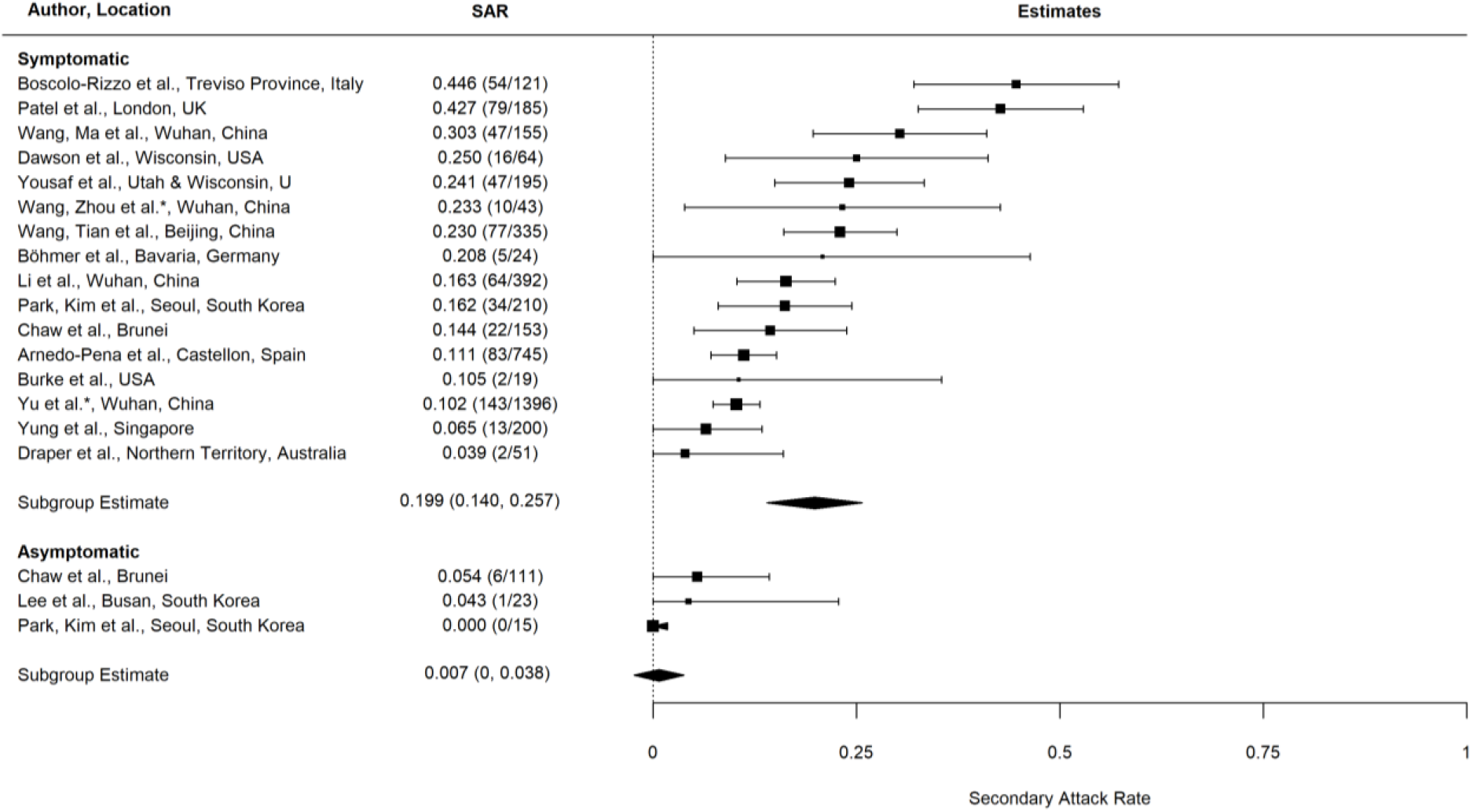
Secondary attack rates (SAR) from symptomatic and asymptomatic index cases to household and family (*) contacts.

Several studies reported SARS-CoV-2 transmission prior to symptom onset of the index case.^22,25,30,52^ One study reported higher risk of transmission after symptom onset relative to the incubation period.^30^ Others demonstrated that exposure after illness onset was not associated with secondary transmission,^26,31,52^ although close contacts and/or index cases were quarantined/isolated after symptom onset. Some studies reported that infection risk peaked during exposure 3–4 days before or within 5 days of symptom onset.^30,52^ One study reported higher SAR among close contacts of presymptomatic index cases than asymptomatic carriers.^52^ Thus, the evidence was mixed.

### Overdispersion

There is evidence for strong clustering of SARS-CoV-2 infections within households, with some households having many secondary infections while many others have none.^56-58^ For example, one study reported that 26 of 103 (25·2%) households had all members test positive.^41^ In China, 78-85% of COVID-19 clusters occurred in families.^11^ Most studies only reported the number of secondary infections per individual, but some also reported transmission by household.^26,28,41,51^ Figure 4 summarizes the proportion of households with any secondary transmission. Using a crude analysis based on the SAR and average number of contacts per household, we find that the proportion of households with any secondary transmission was lower than expected for all studies (see Appendix). Given the full data, fitting a beta-binomial to household data would allow better detection of overdispersion.

**Figure 4.**
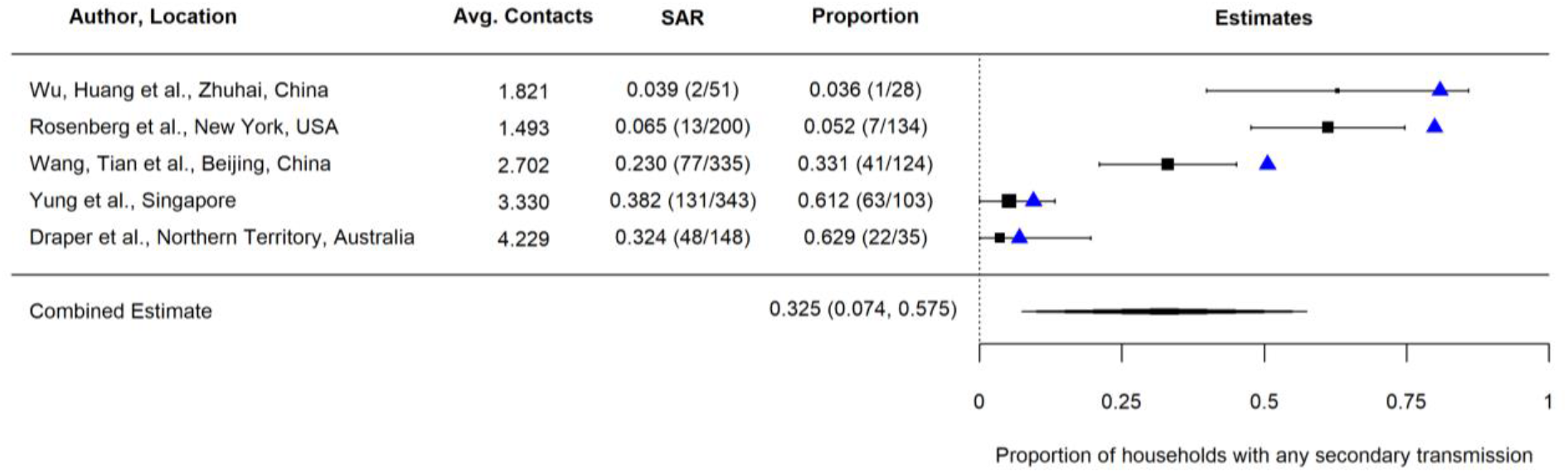
The average number of contacts per household, secondary attack rate (SAR), and proportion of households reporting any secondary transmission from index cases. The blue triangles represent the expected proportion of households with any secondary transmission (see Appendix for further details).

### Risk factors for susceptibility

A number of studies examined factors potentially associated with susceptibility of household contacts to infection (S2 Table). Age was the most examined covariate, with most studies reporting significantly lower secondary transmission of SARS-CoV-2 to children contacts than adult contacts.^16,17,21,22,26,28,30,31,41,43,49^ In three studies, individuals aged >60 years were most susceptible to SARS-CoV-2 infection.^17,22,43^ Contact age was not associated with susceptibility in six other studies,^14,15,24,25,46,52^ although these were typically less powered to detect a difference. Figure 5 summarizes ten studies reporting separate SARs to children and adult contacts. The estimated mean household SAR was significantly higher to adult contacts (31·0%; 95% CI: 19·4%–42·7%) than to children contacts (15·7%; 95% CI: 9·9%–21·5%) (*P* < 0·001), both with significant heterogeneity (*P* < 0·001).

**Figure 5.**
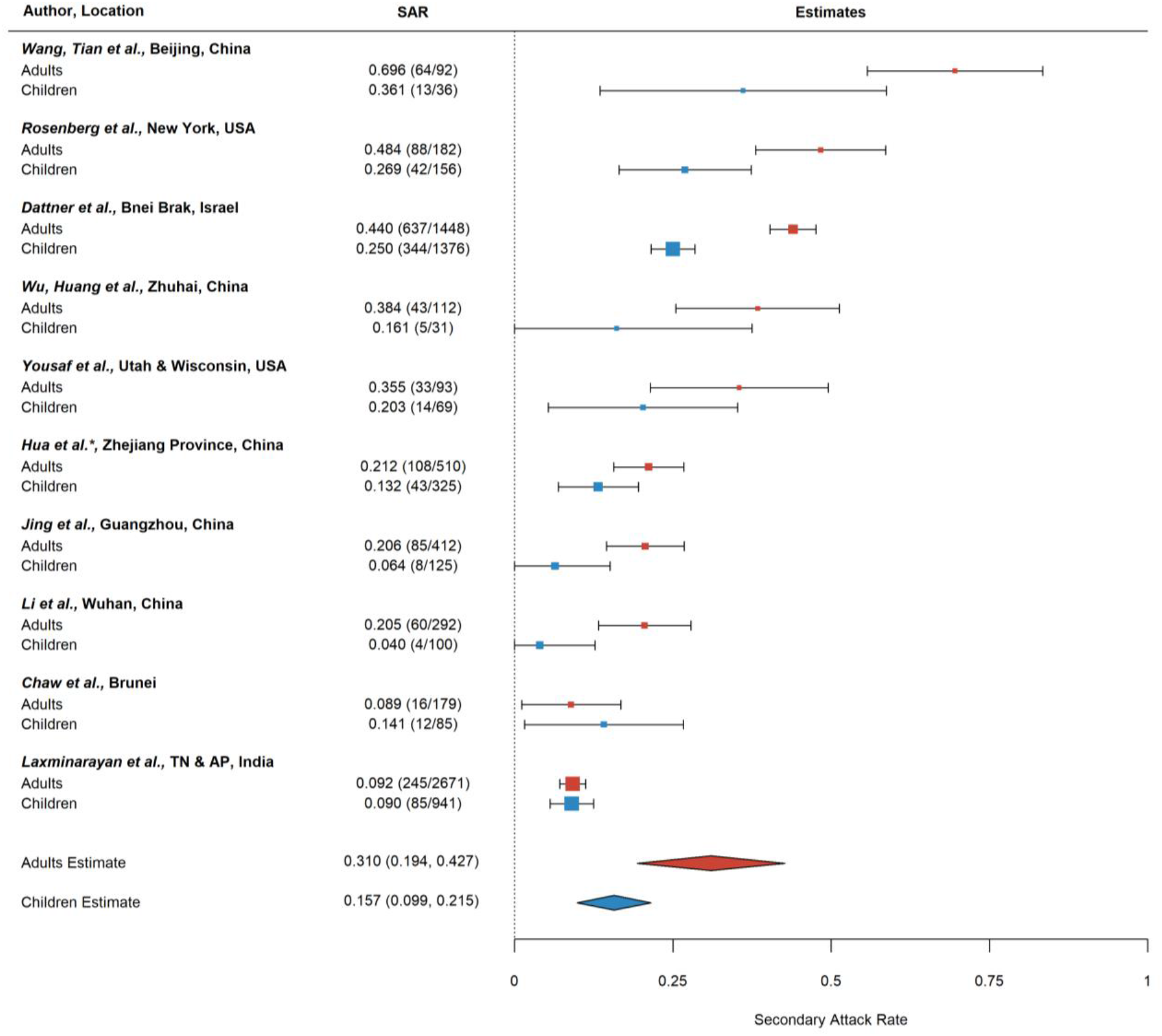
Secondary attack rates (SAR) for adult (≥18 years) and children (<18 years) household and family (*) contacts.

The second most examined factor was sex of the exposed contacts, which was not associated with transmission for most studies,^15,17,18,22,24-26,31,46^ except for two,^16,30^ which found higher risk in females than males. Figure 6 summarizes the results from nine studies reporting household SAR by contact sex. The estimated mean household SAR to females (19·4%; 95% CI: 13·0%–25·8%) was not significantly different than to males (16·0%, 95% CI: 10·3%–21·8%), both with significant heterogeneity (*P* < 0·001).

**Figure 6.**
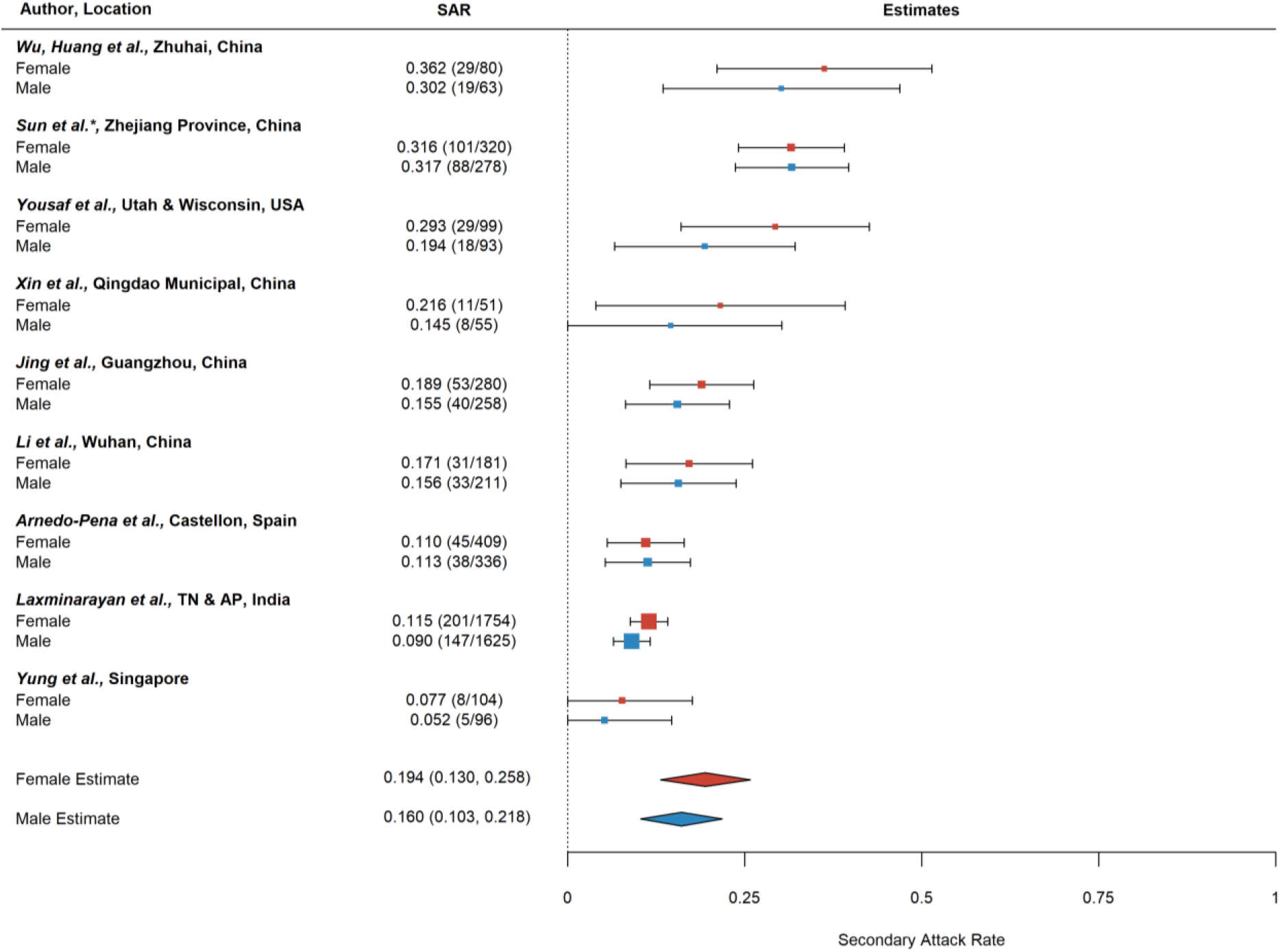
Secondary attack rates (SAR) for household and family (*) contacts by contact sex.

Spouse relationship to index case was associated with secondary infection in four,^15,30,31,46^ of six studies.^14,26^ The risk of infection was highest for spouses, followed by non-spouse family members, close relatives, and other relatives, which were all higher than other contacts.^30^ Figure 7 summarizes the results from five studies reporting household SAR by relationship. The estimated mean household SAR to spouses (43·4%; 95% CI: 27·1%–59·6%) was significantly higher than to other relationships (18·3%, 95% CI: 10·4%–26·2%). There was significant heterogeneity for spouse SARs between studies (*P* = 0·001), but not other relationships. A limitation is that studies do not report the direction of spousal transmission for male-female partnerships.

**Figure 7.**
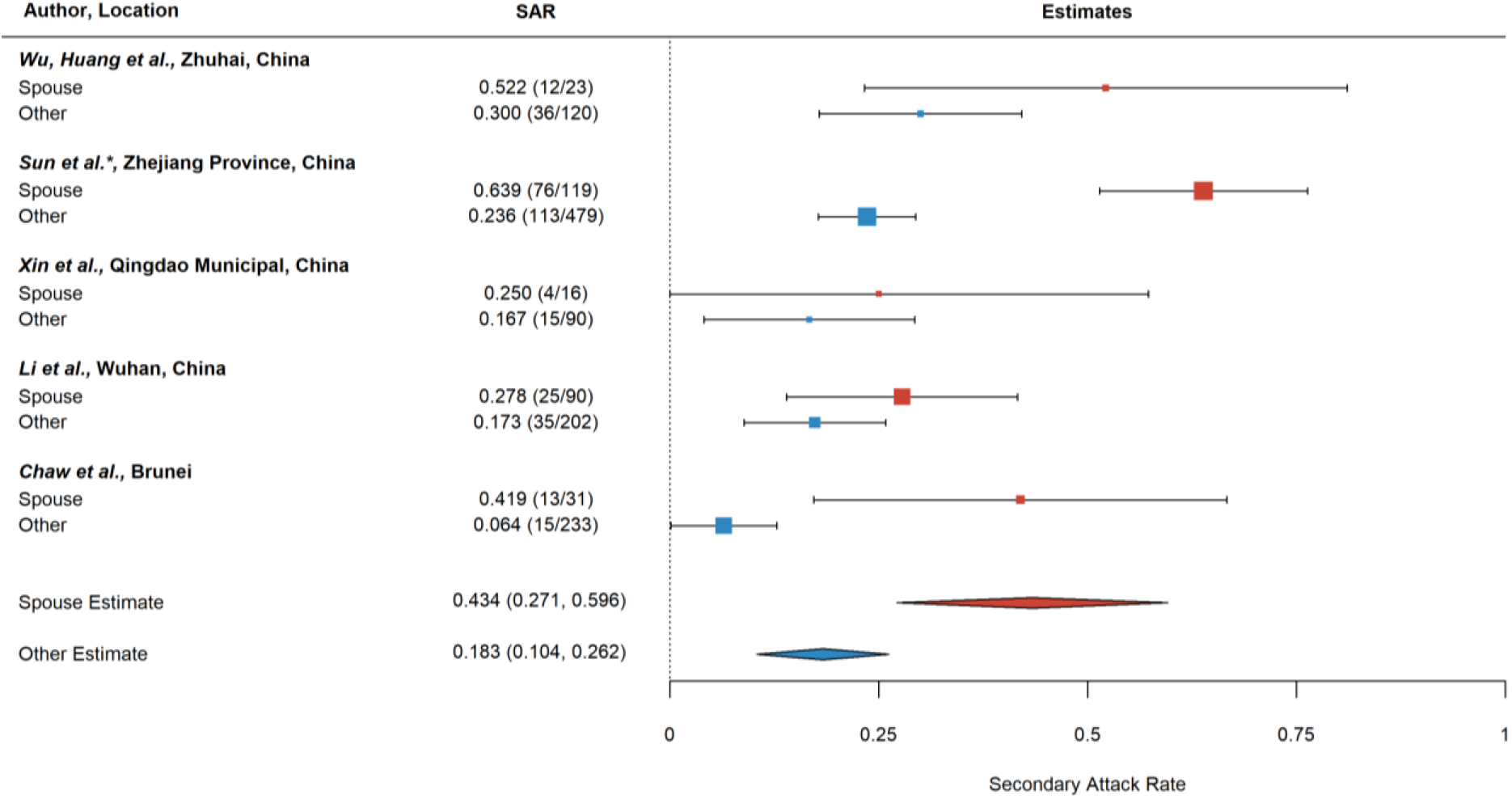
Secondary attack rates (SAR) for household and family (*) contacts by relationship to index case (spouse, other).

### Factors for infectiousness

Several studies also examined factors associated with infectiousness of index cases. Older index case age was associated with increased secondary infections in two,^14,43^ of six studies.^18,22,26,28^ Female index case sex was associated with transmission in two,^14,48^ of six studies.^26,28,31,43^ Figure 8 summarizes the results from six studies reporting household SAR by index case sex. The estimated mean household SAR from females (14·6%; 95% CI: 9·3%–20·0%) was not significantly different than from males (12·8%, 95% CI: 8·1%–17·4%).

**Figure 8.**
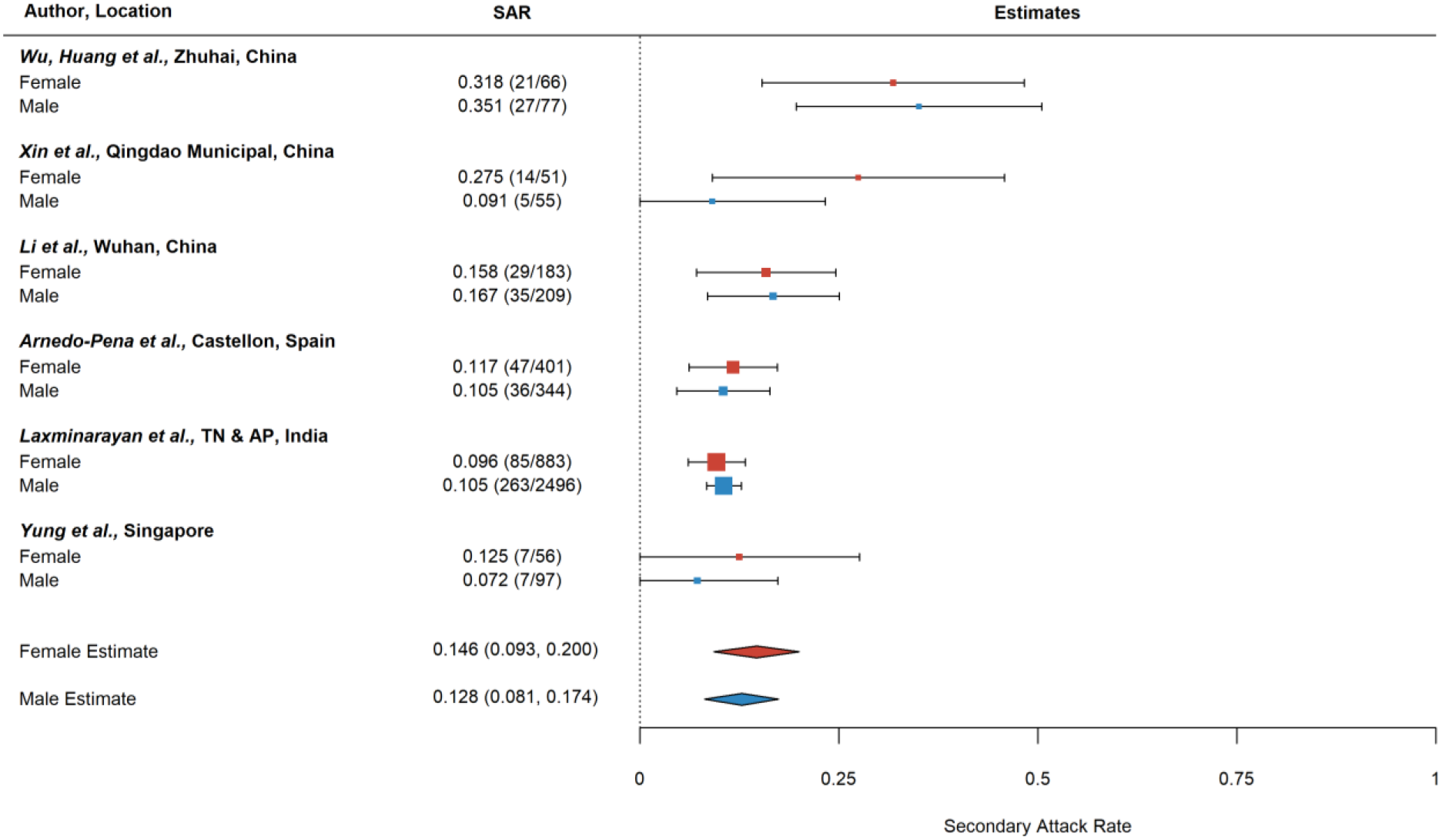
Secondary attack rates (SAR) for household contacts by index case sex.

Critically severe index case symptoms and hospitalization were associated with higher infectiousness in four,^14,17,30,43^ of six studies where this was examined.^25,28^ Cough in the index case was associated with infectivity in two,^26,43^ of seven studies.^14,17,28,30,31^ Diarrhea,^28^ pneumonia,^43,52^ acute respiratory distress syndrome,^52^ myalgia,^30^ chills,^30^ dizziness,^30^ lymphocyte count,^14^ neutrophil percentage,^14^ and expectoration,^17^ were also associated with secondary transmission in some studies. Symptoms not shown to be associated with infectivity were fever,^14,17,26,28,30,31^ fatigue,^14,17,30^ dyspnea,^30^ headache,^30^ nasal congestion,^30^ pharyngalgia,^30^ arthralgia,^30^ rhinorrhea,^30^ nausea,^30^ vomiting,^30^ chest tightness,^30^ palpitation,^30^ poor appetite,^30^ abdominal pain,^30^ and white blood cell count.^14^

Household seroprevalence surveys that did not differentiate correlates of susceptibility vs. infectivity showed similar trends: sex was not associated with infection,^42,56,59,60^ and seroprevalence increased with age.^42,59^

### Awareness and behavioral factors

Contact frequency with the index case was associated with higher odds of infection, specifically ≥5 contacts during 2 days before the index case was confirmed,^25^ ≥4 and 1–3 contacts (within 1 m),^28^ or frequent contact.^14,16,18^ Physical contact,^26^ sharing a vehicle,^17,18,24,26^ sharing a living room,^26^ and sharing a meal,^26,28^ were also associated with infection, but eating with separate tableware was not.^28^ Smoking behavior in index cases or contacts was not associated with transmission.^26^

Several studies explored whether prevention measures were associated with reduced transmission. Contacts who wore face masks and index cases who wore masks all the time after illness onset had lower odds of infection and transmission, respectively.^24,28^ Conversely, contacts who did not apply protective measures (e.g., face mask, avoiding contact with index case) had higher odds of infection.^26^ One study reported that greater frequency of chlorine or ethanol based disinfectant use for house cleaning and ventilation hours per day were associated with reduced risk,^28^ whereas another did not.^26^ Frequency of room cleaning (wet type) and hand hygiene were not significant.^28^ Index case isolation after illness onset was associated with reduced secondary transmission.^31^ Other studies did not find the time interval from illness onset to medical isolation,^28^ hospital admission,^14,26,28^ or laboratory confirmation,^14,28^ to be associated with transmission. Self-awareness of being infected with SARS-CoV-2 and knowledge of COVID-19 were not significant,^28^ but lack of knowledge of index case’s own infectiousness was associated with transmission.^28^ Health profession of the index case was a protective factor in one study.^43^

### Household characteristics

Smaller households were associated with transmission in two,^22,43^ of six studies.^26,28,41,42^ Figure 9 summarizes the results from four studies reporting household SAR by number of contacts in the household. The estimated mean household SAR for households with one contact (45·2%; 95% CI: 34·1%–51·8%) was significantly higher than households with ≥three contacts (25·1%; 95% CI: 11·1%– 39·1%) (*P* < 0·001), but not significantly different than households with two contacts (47·0%; 95% CI: 16·8%–77·1%). There was significant heterogeneity in SAR between studies with two (*P* < 0·001) or ≥three contacts (*P* < 0·001), and but not one contact. A limitation here is that we have no information about household crowding (e.g., number of people per room).

**Figure 9.**
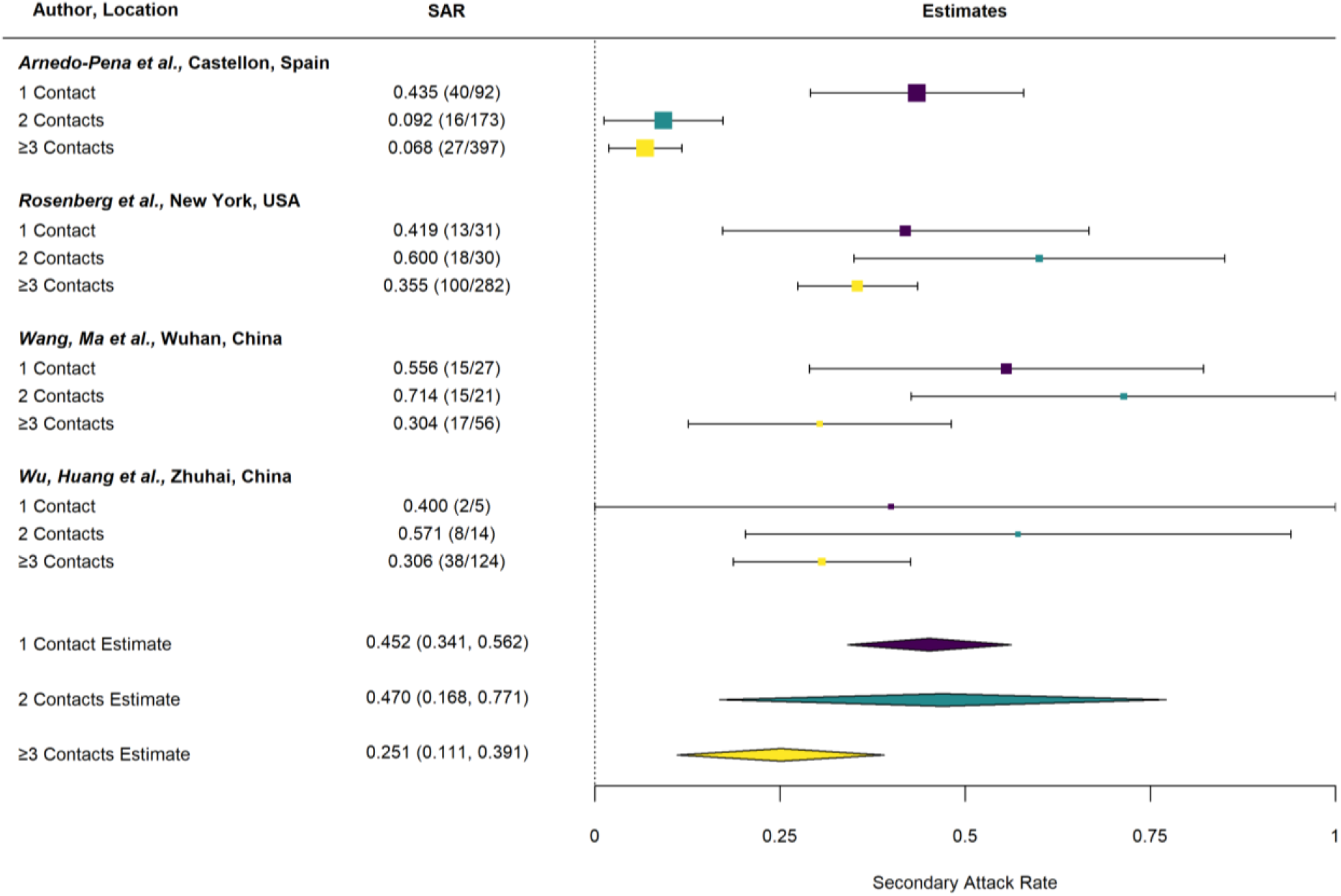
Secondary attack rates (SAR) by the number of contacts in the household.

### Comparison with other viruses

To compare SARS-CoV-2 SARs with other viruses, we reviewed articles describing household secondary transmission of SARS-CoV, Middle East Respiratory Syndrome coronavirus (MERS-CoV), and other coronaviruses. We found seven articles reporting household SARs of MERS-CoV,^61-67^ six of SARS-CoV,^68-73^ and four of other coronaviruses (S3 Table).^74-77^

The estimated mean household SAR was 6·0% (95% CI: 2·2%–9·8%) for SARS-CoV with no significant heterogeneity and 3·5% (95% CI: 0·1%–6·8%) for MERS-CoV with no significant heterogeneity (Figure 10), which were both lower than the overall household SAR of 18·8% for SARS-CoV-2 in this study (*P* = 0·002). The SAR for SARS-CoV-2 was also higher than SARs reported for other coronaviruses: 0–12·6% for HCoV-NL63, 10·6–13·2% for HCoV-OC43, 7·2–14·9% for HCoV-229E, 8·6% for HCoV-HKU1,^74,76,77^ and 2·6% for an unspecified novel coronavirus.^75^ Household SARs for SARS-CoV-2 appear to be within the upper ranges of household SARs reported for influenza, which ranged from 1–38% based on PCR-confirmed infection, 6–35% based on influenza-like illness, and 3– 31% based on acute respiratory illness.^13^

**Figure 10.**
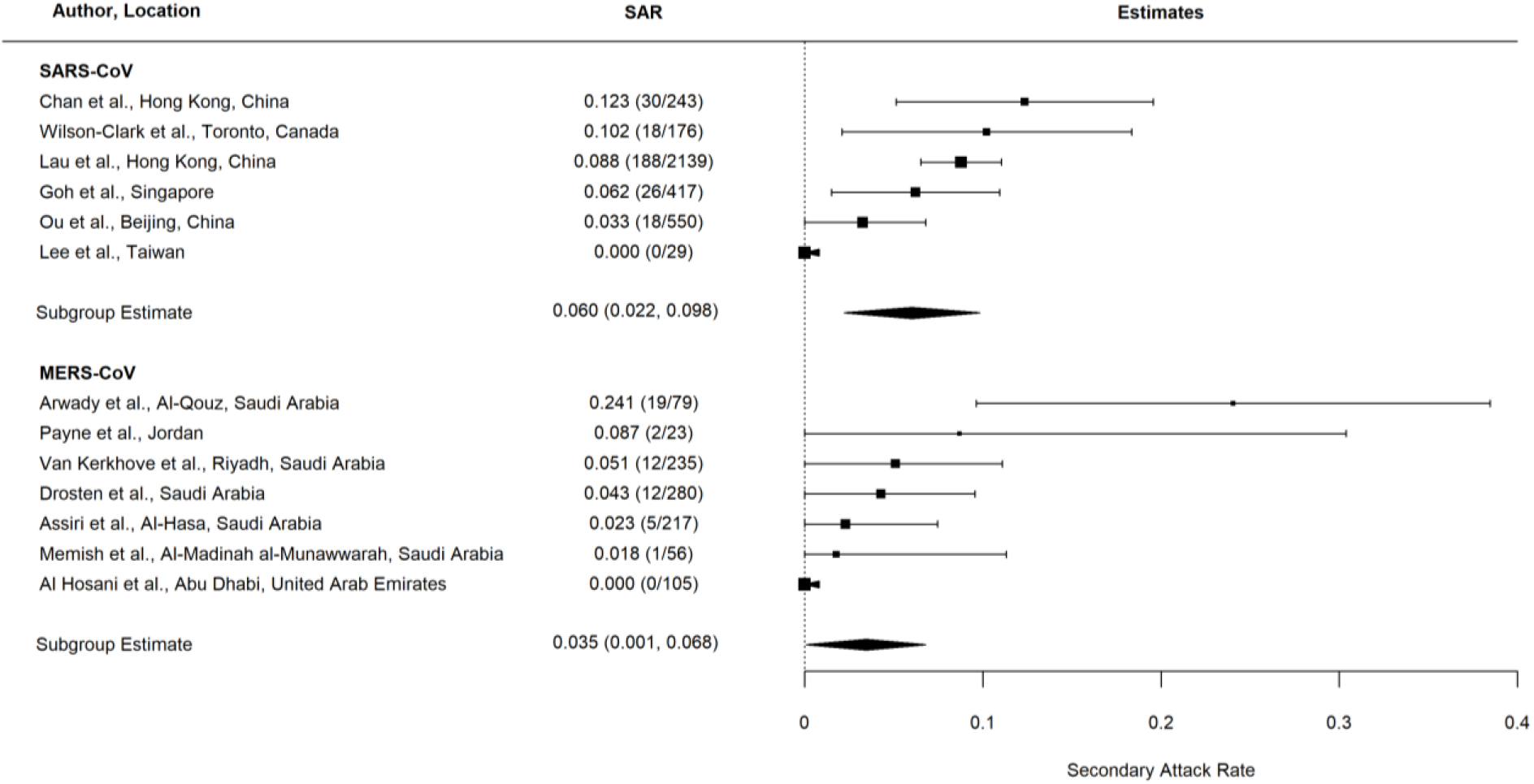
Household secondary attack rates (SAR) for severe acute respiratory syndrome coronavirus (SARS-CoV) and Middle East Respiratory Syndrome coronavirus (MERS-CoV).

## Discussion

We synthesize available evidence on household studies of SARS-CoV-2. Combined household and family SAR is approximately 19%, which is higher than previously observed SARs for SARS-CoV and MERS-CoV. Results suggest that household and family contacts are at higher risk than other types of contacts. We observed that household SARs were higher from symptomatic index cases than asymptomatic index cases, to adult contacts than children contacts, to spouses than other family contacts, and in households with one contact than households with three or more contacts. We also found some limited evidence of overdispersion in the number of infections caused by index cases, highlighting potential heterogeneity in transmissibility of index cases.

The household may be a favorable environment for transmission due to the frequency of contacts between family members, reduced usage of personal protective equipment, shared living and eating environment, persistence of SARS-CoV-2 on different surfaces,^78^ and potential fecal shedding in shared toilets.^79^ Modeling studies demonstrated that household transmission had a greater impact on *R* after social distancing (30%-55%) compared to before social distancing (5%-35%).^80^

We found significantly higher SARs from symptomatic index cases than asymptomatic index cases, but many studies acknowledged potential underreporting of asymptomatic cases. There were also relatively few studies with separately reported asymptomatic index cases. Prolonged unprotected exposure to symptomatic case patients increases risk of transmission through respiratory droplets, by direct contact, or contact with fomites.^11^ Modeling studies have suggested, however, that asymptomatic index cases may also be important drivers of SARS-CoV-2 transmission, particularly from presymptomatic cases.^81-84^

SARs were higher to adult contacts than children contacts. Lower infection rates in children may in part be attributed to asymptomatic or mild disease and low case ascertainment.^85^ Few household studies reported SARs by index case age, but data suggests children have not played a substantive role in household transmission of SARS-CoV-2.^86-88^ A large study in South Korea that included 10,592 household contacts, however, noted relatively high transmission from index cases who were 10–19 years of age contracted COVID-19.^33^ Although children seem to be at reduced risk for symptomatic disease, it is still unclear whether they shed virus similar to adults.^89^

The finding that households with only one contact had higher SARs than households with greater than two contacts may suggest that household crowding and residential area per person are more important for SARS-CoV-2 transmission than the total number of people per household. Household crowding (e.g., the number of people per room) was demonstrated to be associated with influenza transmission.^90-92^

The observation that spouse relationship to the index case was a significant risk factor for secondary transmission is consistent with studies of SARS-CoV and H1N1.^93,94^ This may be a reflection of intimacy, sleeping in the same room, or longer or more direct exposure to index cases. One study detected SARS-CoV-2 in semen.^95^ Further investigation is required to determine whether sexual contact is a transmission route.

We did not find associations between household contact or index case sex and secondary transmission. WHO reports roughly even distribution of SARS-CoV-2 infections between women and men worldwide with higher mortality in men,^96^ perhaps due to delayed viral clearance of SARS-CoV-2,^97^ but acknowledges limited availability of sex-aggregated data. One study of mice showed that female mice were less susceptible to SARS-CoV than male mice, due in part to protection from estrogen receptor signaling and activity of X-linked genes.^98^

Household SARs were higher for SARS-CoV-2 than SARS-CoV and MERS-CoV, which may be attributed to structural differences in spike proteins,^99^ higher basic reproductive rates,^100^ and higher viral loads in the nose and throat at the time of symptom onset.^101^ Severe symptoms associated with MERS and SARS often require hospitalization, which increases nosocomial transmission, whereas less severe symptoms of SARS-CoV-2 facilitate community transmission.^101^

Although the pandemic transcends languages and culture, standardization of reporting procedures would facilitate comparisons of published studies. To expedite comparison of studies, it would be helpful for investigators to report the number of infected contacts and total contacts by specific characteristics of contacts (e.g., age group, sex, relationship, others) and by characteristics of index cases (e.g., symptomatic/asymptomatic, age group, others). To better understand clustering within households, it would also be useful for researchers to report the number of infections by household in addition to the total number of infected individuals.

Our study had several limitations. The large amount of unexplained heterogeneity across studies could be attributable to variability in study definitions of index cases and household contacts, frequency and type of testing, sociodemographic factors, household characteristics (e.g., density, air ventilation), rates of community transmission, and local policies (e.g., centralized isolation). Many of the studies involved testing symptomatic household contacts, which likely missed asymptomatic infections, although SAR estimates were similar across studies testing all contacts and only symptomatic contacts.

Conversely, the estimates may overestimate household transmission from index cases to contacts because studies cannot typically rule out infection from outside the home (e.g., non-household contact, fomite).

Important questions remain about the household spread of SARS-CoV-2 including the efficiency of asymptomatic transmission, probability of fecal-oral transmission, role of children in potential of reinfection, and sexual transmission of SARS-CoV-2.^102^ To prevent the spread of SARS-CoV-2, people are being asked to stay at home worldwide. With suspected cases frequently referred to isolate at home, household transmission will continue to be a significant source of transmission. Prevention strategies such as increased mask-wearing in the home, improved ventilation, voluntary isolation at external facilities, and targeted antiviral prophylaxis should be explored.

## Data Availability

All relevant data are within the manuscript.

## Declaration of interests

We declare no competing interests.

## Acknowledgments

This work was supported by the National Institutes of Health R01-AI139761.

## Notes

### Competing Interest Statement

The authors have declared no competing interest.

